# Labelling chest x-ray reports using an open-source NLP and ML tool for text data binary classification

**DOI:** 10.1101/19012518

**Authors:** Sohrab Towfighi, Arnav Agarwal, Denise Y. F. Mak, Amol Verma

## Abstract

The chest x-ray is a commonly requested diagnostic test on internal medicine wards which can diagnose many acute pathologies needing intervention. We developed a natural language processing (NLP) and machine learning (ML) model to identify the presence of opacities or endotracheal intubation on chest x-rays using only the radiology report. This a preliminary report of our work and findings. Using the General Medicine Inpatient Initiative (GEMINI) dataset, housing inpatient clinical and administrative data from 7 major hospitals, we retrieved 1000 plain film radiology reports which were classified according to 4 labels by an internal medicine resident. NLP/ML models were developed to identify the following on the radiograph reports: the report is that of a chest x-ray, there is definite absence of an opacity, there is definite presence of an opacity, the report is a follow-up report with minimal details in its text, and there is an endotracheal tube in place. Our NLP/ML model development methodology included a random search of either TF-IDF or bag-of-words for vectorization along with random search of various ML models. Our Python programming scripts were made publicly available on GitHub to allow other parties to train models using their own text data. 100 randomly generated ML pipelines were compared using 10-fold cross validation on 75% of the data, while 25% of the data was left out for generalizability testing. With respect to the question of whether a chest x-ray definitely lacks an opacity, the model’s performance metrics were accuracy of 0.84, precision of 0.94, recall of 0.81, and receiver operating characteristic area under curve of 0.86. Model performance was worse when trained against a highly imbalanced dataset despite the use of an advanced oversampling technique.

## Background

The chest x-ray (CXR) is the most commonly requested imaging test and is critical for the diagnosis of several conditions, including pulmonary infections [1]. The CXR has been found to provide valuable clinical data, often supporting the clinically suspected diagnosis, identifying previously unknown pathology, and influencing the course of clinical management [2]. Given its clinical utility and the insight it provides into the patient’s medical state, the CXR report is of value to researchers performing retrospective studies. Being able to automatically determine whether CXR reports identify a specific pathology permits researchers to identify members of a cohort for further study.

We are interested in analysing CXR reports from the General Internal Medicine Inpatient Initiative (GEMINI) database, a multicentre dataset which links administrative and clinical data across 7 hospitals affiliated with the University of Toronto [3]. We are particularly interested in two characteristics that can be determined from CXR reports: whether there is the presence of an opacity that could be consistent with pneumonia and whether the patient has an endotracheal tube in place. Pneumonia is one of the most common causes of hospital admission, representing 5% of admissions to General Internal Medicine [4]. Pneumonias are associated with significant inpatient hospital admission time, morbidity and mortality. Intensive care and intubation may be involved in the trajectory of these patients [5]. Large-scale health databases of clinical data include CXR reports that provide evidence of infiltrate and possible intubation. However, without labeling of radiographic reports as having or not having these properties, the use of these reports in large-scale research analyses may be limited. Developing automated methods of identifying pneumonia in clinical datasets could substantially improve the reliability of detecting this important condition in health services research and quality measurement, as compared to typical methods based on administrative diagnostic codes. Similarly, endotracheal intubation is a key intervention for patients with critical illness and is associated with pneumonia in a small proportion of cases. Automating a method to reliably identify whether patients have developed ventilator-associated pneumonia could also be useful for research and quality improvement.

Classifying radiology reports based on presence of opacity or endotracheal tube is an important step toward identifying pneumonia and ventilator-associated pneumonia in clinical datasets.

ML approaches have been applied for the extraction of key data from medical notes and even more challenging tasks like directly interpreting medical images [6]. ML algorithms solve either regression or classification problems, and function in a supervised or unsupervised manner. Supervised classification problems have a ground truth dataset against which the model is trained, whereas unsupervised algorithms segregate the data into clusters without any a priori knowledge about the true classes. ML algorithms appear on a spectrum with respect to their performance and interpretability. The most successful ML algorithms for image interpretation have been Deep Learning (DL) algorithms. DL models are a network of data manipulating operators that work parsimoniously to solve classification and regression supervised learning problems. This network is usually too complex to be interpretable and so is viewed as a black box by most users [6].

NLP encompasses various computing approaches to using free-form text data in analyses [7]. When applied to medical data, NLP can convert text data into a structured and numerical form more accessible by ML algorithms. It can also serve to distill redundancies in how we form language, which improves the result of subsequent analyses. NLP has many applications in radiology, including identifying pneumonia, pulmonary nodules, and even BI-RADS scores from radiology reports. Rule-based systems, where expert domain knowledge is used to handcraft a set of rules that specify how to classify text, are an alternative to ML for text classification problems. Rule-based approaches can lead to accurate models, but they require more work to develop and may not generalize as well as statistical ML models [7].

The objective of this study was to develop an automated tool to classify radiology reports to identify opacity and/or endotracheal intubation, which could be associated with pneumonia. We use data from 7 hospitals in Ontario, Canada, collected through the GEMINI project, to enhance the generalizability of our tool. Our approach uses machine learning (ML) and natural language processing (NLP) based on open-source software and a custom-developed pipeline. Our ML analyses use popular and state-of-the-art Python codes which we provide as a command line interface script, housed on GitHub. This document is a preliminary report of our work and findings.

## Method

### Design and Setting

The General Medicine Inpatient Initiative (GEMINI) study is a retrospective cohort study including 7 hospitals affiliated with the University of Toronto. Participating institutions include St. Michael’s Hospital, Mount Sinai Hospital, Sunnybrook Health Sciences Centre, Trillium Health Partners (Credit Valley and Mississauga sites) and the University Health Network (Toronto General Hospital and Toronto Western Hospital). Each health care organization participating in GEMINI is independent. The GEMINI hospitals are academic teaching hospitals where medical residents and students rotate. Data were retrieved from the GEMINI database and included patients having an x-ray report during a hospital admission from 2010-2017 [3].

Patients with an empty x-ray report were excluded and one x-ray report was randomly chosen for patients having multiple x-ray tests during their admission. A total of 1000 x-ray reports were randomly selected for inclusion.

### Labelling Chest X-ray Reports

The imaging reports were classified by an internal medicine resident with respect to the following variables.

1. The report is that of a CXR
2. The report definitely has no opacity suggesting pneumonia
3. The report definitely has an opacity suggesting pneumonia
4. The report is of a follow-up imaging report with minimal details in the text of the report
5. The report shows the presence of an endotracheal tube

The use of definitive language in formulating our questions implies that cases where there is uncertainty are relegated to being classified as false. Thus, the classification based on items 2 and 3 are not perfect complements of each other, but rather uncertain cases are categorized differently in the two approaches. In many instances, CXR reports in hospital are simply reported as “no change from previous”, particularly if radiographs are being performed over consecutive days for the same patient. Thus, classification task 4 would allow the identification of these reports.

### Natural Language Processing

We performed tokenization using the spacy [8] Python package. We transformed all text into lower case, and removed punctuation, English language stop words, and personal pronouns. After this preprocessing, the data was passed into one of two vectorizers. One of our vectorizers was a Bag-of-Words vectorizer which counted the number of times a word was found in a document and converted each x-ray report into a numerical vector. Since all the words in our corpus would be represented within this vector, each document was converted into a vector that is mostly zeros, also known as a sparse vector. As there are combinations of words that may be valuable features, we also employ the N-gram paradigm to consider pairs or triplets of words as permitted features. Interested readers are referred to the following reference for further information regarding the Bag-of-Words representation of text data [9]. Our second vectorizer was the Bag-of-Words representation with an added step of applying a term-frequency times inverse document-frequency (TF-IDF) transformer. The TF-IDF transform takes the previously described sparse vector, and multiplies the value of each element (or term) by the frequency of the term in the entire set of documents multiplied by the logarithm of the quotient of the total number of documents and the number of documents in which the term appears. Practically speaking, the TF-IDF transformation decreases the weight of terms which appear in most documents and increases the weight of terms found in a small set of documents. Readers interested in a thorough discussion of the historical origins and theoretical arguments for the TF-IDF transform are referred to the following reference [10].

### Machine Learning

After the data has been passed through the NLP including vectorization operations, we perform additional preprocessing prior to ML. For datasets where one class is much more common than the other, ML classification algorithms may generate models with poor generalizability. The algorithms may produce models which always predict that a data point belongs to the more common class. In order to avoid this issue, we pass the training data to a synthetic minority over-sampler [11] which generates artificial data points belonging to the less commonly occurring class. The test dataset has no oversampling performed on it. After this oversampling step, the data is ready to be considered by the ML algorithms. Our model selection is entirely random, so a random text vectorizer is generated along with a random ML algorithm to produce a random pipeline.

The ML models we consider are ones included in the very popular scikit-learn library [12]. These models include random forests, gradient boosting, linear support vector, logistic regression, and neural networks. Each ML algorithm produces a predictive model with its own parameters and the performance of the resultant model is dependent on the hyperparameters used by the algorithm during its run. The choice of hyperparameters is a complex topic, but recently, it was found that random search for hyperparameter selection is a competitive approach [13], so we apply this methodology for our hyperparameter search and extend its use to model selection. The values of the hyperparameters through which we randomly search were taken from the source code of the popular automated ML pipeline generator, TPOT [14]. We randomly generate a vectorizer, an ML algorithm, its hyperparameters, and then we fit this randomly generated pipeline to the training dataset.

The model along with its performance metrics are recorded in a SQLite database file. SQLite is a nimble relational database system housed within a single file which does not require a separate server. We use a Python package for SQLite which works like a dictionary, enforcing uniqueness of keys. This setup ensures that if the random pipeline generator happens to produce a previously considered model, duplicate results are not saved. The benefit of this approach to model selection in contrast to a grid search of different ML algorithms is that grid-search is computationally expensive and is better suited when we are interested in separating algorithms and comparing the best performing configuration of an algorithm with the best performing configuration of another algorithm.

Since we are working with algorithms that have been extensively studied, and our goal is simply to generate an accurate classifier in a timely manner, we use random search. In supervised ML analysis, the data is split into a training set, on which the model is trained, and a testing set, on which the generalizability of the model is tested. In our case, we split the data such that we train on 75% of the data and test on 25% of the data. Cases were ordered randomly. For our model selection using the training data, we perform a 10-fold cross validation to calculate performance metrics which are used in ranking the quality of the models. Interested readers are referred to the following work, in which cross-validation strategies are explored [15].

### Open Source Script

Our software is available to interested readers in an open-source GitHub repository: github.com/pySRURGS/nlp_ml. The code is a command line interface script which takes a few key arguments. The user supplies the script with file paths to comma separated value (CSV) files housing the training and testing datasets that the user must prepare beforehand, the number of different pipelines to consider, and the file path to where the output database file should be saved. The software was written in Python version 3.6 and is released with an open source license, the GPL version 3.0 license.

## Results

There were 208 x-ray reports that were not CXR reports in our labelled dataset (despite being categorized as CXR based on hospital test naming conventions), which leaves 792 reports for questions 2-5. The error matrices generated for the datasets are plotted in Figure 1, and the receiver-operating-curve (ROC) plots are found in Figure 2. The results of each labelling task are summarized in Table 1.

**Table 1:** Performance metrics of the best models for each of the five questions^1^

| Question | Training Accuracy | Training Precision | Training Recall | Train ROC AUC | Test Accuracy | Test Precision | Test Recall | Test ROC AUC |
| --- | --- | --- | --- | --- | --- | --- | --- | --- |
| 1 | 0.99 | 1 | 0.98 | 0.99 | 0.98 | 0.98 | 0.98 | 0.97 |
| 2 | 0.89 | 0.92 | 0.86 | 0.89 | 0.84 | 0.94 | 0.81 | 0.86 |
| 3 | 0.98 | 0.96 | 1 | 0.98 | 0.87 | 0.38 | 0.4 | 0.66 |
| 4 | 1 | 1 | 1 | 1 | 0.96 | 0.25 | 0.2 | 0.59 |
| 5 | 1 | 1 | 1 | 1 | 1 | 1 | 1 | 1 |

**Figure 1:**
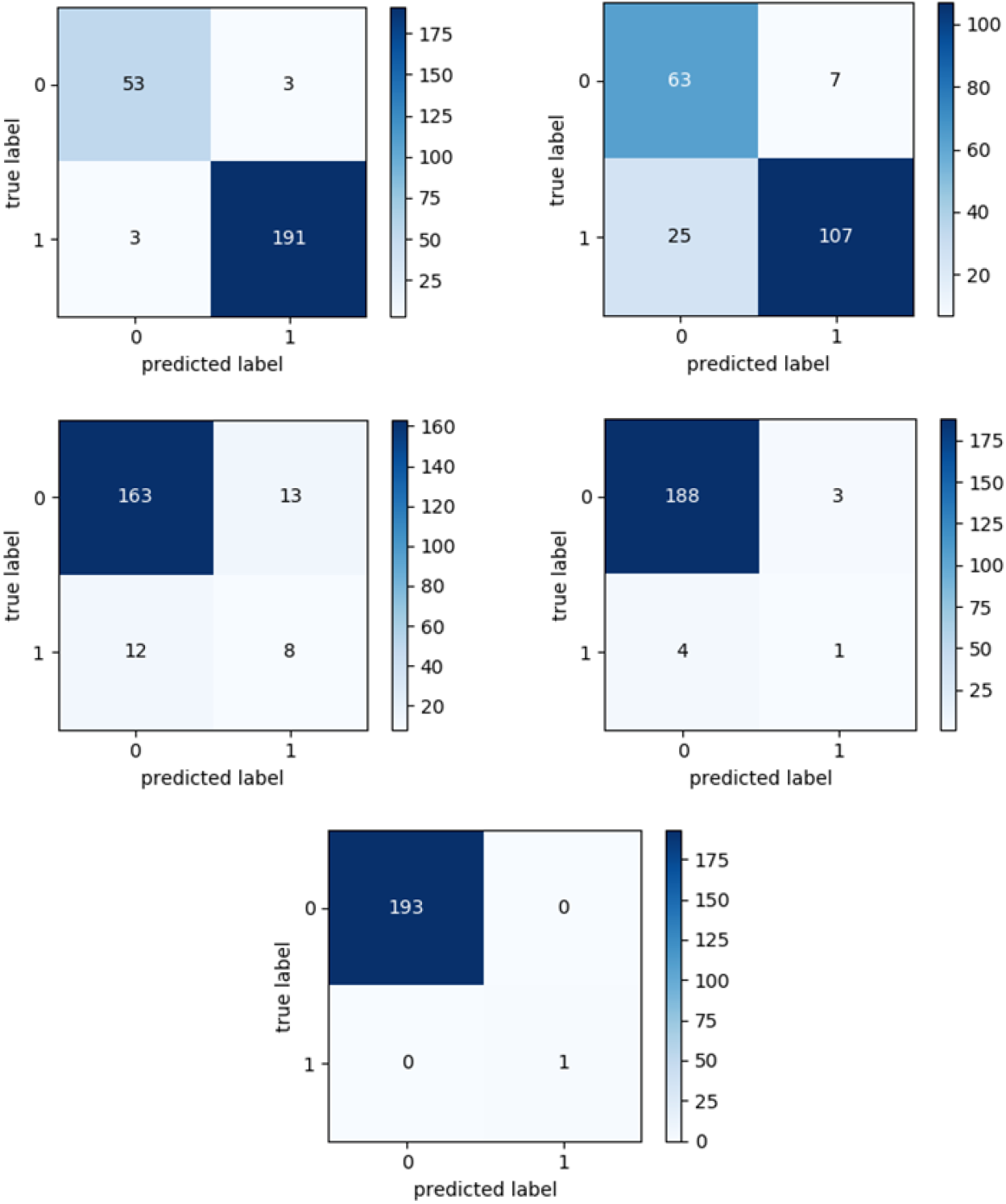
Error matrices of the testing dataset for the five questions being considered. Top left corresponds to (1. the report is that of a CXR), top right to (2. the report definitely has no opacity suggesting pneumonia), middle left to (3. the report definitely has an opacity suggesting pneumonia), middle right to (4. the report is of a follow-up imaging report with minimal details in the text of the report), and bottom to (5. the report shows the presence of an endotracheal tube).

**Figure 2:**
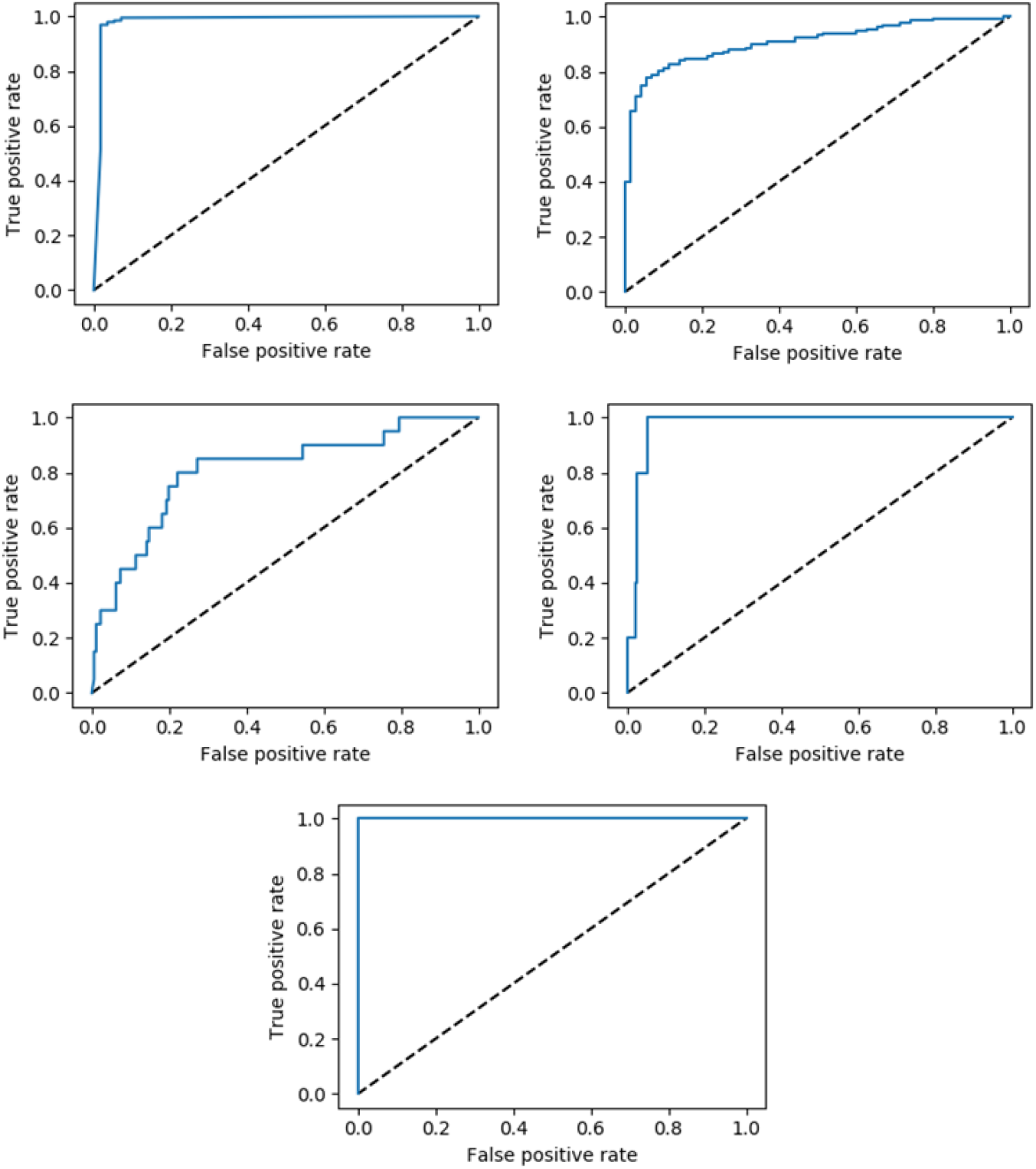
Receiver Operating Characteristic plots of the testing dataset for the five questions being considered. Top left corresponds to (1. the report is that of a CXR), top right to (2. the report definitely has no opacity suggesting pneumonia), middle left to (3. the report definitely has an opacity suggesting pneumonia), middle right to (4. the report is of a follow-up imaging report with minimal details in the text of the report), and bottom to (5. the report shows the presence of an endotracheal tube).

The model responsible for identifying cases definitely without opacity had an accuracy of 0.84, a precision of 0.94, a recall of 0.81, and a ROC AUC of 0.86.

We consider whether a given radiograph is for a follow-up image, in which the radiologist simply dictates that there is no change from prior. These reports occur infrequently, and our algorithm has a low precision value of 0.38 against the testing data. We also consider whether the radiograph report specifies that the patient was intubated. There was only one instance where the patient was intubated in the test set, and our model performed the classification correctly.

## Discussion

There are studies considering the use of automated methods to identify pneumonia from chest x-ray reports. In an early study [16], workers found that a decision tree model, a rule-based model, and a Bayesian model all performed similarly to physicians in classifying a radiograph’s report as supporting pneumonia. They report that their rule-based system had a sensitivity of 0.92 a precision of 0.80, and a specificity of 0.86. Their Bayesian network classifier had a sensitivity of 0.90, a precision of 0.72 and a specificity of 0.78. Lastly, their decision tree model had a sensitivity of 0.86, a precision of 0.85 and a specificity of 0.91. In their study, the gold standard was the majority vote classification of three physicians. More recent work used a rule-based system to identify definite cases of pneumonia from only CXR report and reported good results [17]. They had a very large training dataset of 93,000 CXR reports, and their testing dataset comprised 5000 CXR reports. Their most accurate model had a sensitivity of 92%, specificity of 87% and precision of 74%. This model excluded 25% of the reports due to uncertainty and including these reports would likely decrease the performance metrics they report. There has also been work done to determine whether pneumonia can be identified from emergency department notes [18], which found acceptable classification results (sensitivity of 0.89 and precision of 0.80) when combining NLP and diagnostic code data prior to the use of a support vector machine ML classifier.

Our study differs in the following respects: we provide our general-purpose NLP/ML code in an open source GitHub repository, and we use a random search for both model selection and hyperparameter optimization. Our first classification task was to determine whether a given report is that of a CXR or not. This is expected to be a relatively simple classification problem because, intuitively, most chest x-ray reports have some configuration of ‘chest’ and ‘x-ray’, so it is not surprising that the machine learning model is effective in its ability to separate CXR reports from other modality radiology reports. The performance metrics of our classifier predicting definite absence of opacity were good, with a high positive predictive value, which makes it useful for our future analysis of the entire GEMINI dataset. Conversely, the performance metrics of our classifier predicting definite presence of opacity were poor, possibly because there were too few cases of pneumonia in the training dataset.

In developing the software, we realized that our dataset was highly imbalanced and required an oversampling algorithm in order to prevent the ML classifier from simply predicting the most common label. Incorporating the SMOTE oversampler makes our software more robust, being able to produce good classifiers even when the dataset has fewer abnormal cases, which is likely to be the case in most medical datasets. As demonstrated by our performance on the definite opacity classification task, the SMOTE oversampler is not able to overcome the degree of imbalance in the data. Our choice of cross-validation methodology is also well considered.

Performing 10-fold cross-validation on the training dataset gives us an unbiased estimate of classifier performance and allows us to select the model with the most generalizable results for production system use and is not as computationally intensive as leave-one-out cross-validation. Since our dataset is relatively large, leaving 25% of the total sample for the test phase is sufficient for a reasonable verification of our estimated generalizability performance.

Since our software is released under an open source license, the GNU Public License Version 3.0 license, interested parties can use it for their own medical database labelling purposes without charge. Since computing resources within hospital infrastructure are difficult to acquire and maintain, it is important that the software be relatively easy to install and require only modest computer hardware. We were able to use minimal computational resources, running our computations against private health data using our institutional library’s computer systems and demonstrating that the software is nimble and practical. The software runs on Windows 10 operating system and was installed without administrator privileges. However, its installation and operation are done from the command line so would require some technical knowledge. For our use, we used the Git Bash terminal when installing and using the software, which permits the use of various Linux command line tools and allows for Bash language syntax when navigating the computer’s folders and running commands. Most data scientists and IT professionals would be familiar with these topics and their assistance can be sought when installing and using the software.

Prior to deploying the models against the GEMINI datasets, we plan to increase the size of our manually labelled dataset, and we expect our performance improvement to be significant, especially with respect to the problem of identifying cases of definite opacity and cases where the report is that of a follow-up without diagnostically informative text. For the datasets which are relatively balanced, the models perform very well against their testing datasets, but for those with highly imbalanced datasets, the models have poorer performance. There exist large datasets of publicly available labelled CXR reports including the CheXpert [19] and MIMIC [20] datasets, which were labelled using an automated rule-based method. The CheXpert dataset reports a ROC AUC ranging between 0.85-0.97 on their testing dataset of 500 studies labelled by board certified radiologists.

Our study has several weaknesses. Since NLP/ML algorithms learn with increasing exposure to positive findings, we are limited by the size of our sample and the frequency of positive findings. We have refrained from using deep learning (DL) algorithms. Ad hoc tests we ran with recurrent neural networks resulted in memory usage in the range of 20 gigabytes of random-access-memory, which is untenable for us. Our study has several strengths. We use open source code, providing the community with a methodology that they can use for their own free-text classification problems. Our models perform very well against training data that is relatively balanced.

This manuscript described a preliminary paper of a multicentre study involving chest x-ray reports from seven hospitals in Ontario, Canada, demonstrates that a set of open-source NLP and ML tools was able to discern distinguishing patterns from the radiograph reports and classify chest x-rays with clinically meaningful labels. Although the general NLP and ML methodology used here is well established, we incorporated an entirely random search of both hyperparameters and algorithm selection. This method was able to identify radiographs as chest x-rays, classify radiographs as having or not having opacity consistent with pneumonia, identify radiographs reported as unchanged follow-ups from previous, and identify the presence of an endotracheal tube. Our preliminary results demonstrate good discrimination across tasks. This tool, when applied to the larger GEMINI dataset (approximately 245,000 hospital admissions), will facilitate a variety of research and quality reporting applications.

## Conclusion

In this preliminary report, we found that our NLP/ML models performed well on supervised classification tasks and generated models that will be of use in annotating much larger datasets from the GEMINI database. With respect to the question of whether a chest x-ray definitely has no opacity, the model’s performance metrics were accuracy of 0.84, precision of 0.94, recall of 0.81, and ROC AUC of 0.86. The performance of generated models deteriorated when trained against highly imbalanced data. The software used in this analysis is released as open-source code, free for researchers to use. We demonstrate that the methodology described herein is effective in tackling both challenging and simpler text classification problems using real-world clinical data from major hospital electronic medical record systems.

## Data Availability

The data is not publicly available.

## Acknowledgements

The General Medicine Inpatient Initiative (GEMINI) was supported by grants from the Green Shield Canada Foundation and the University of Toronto, Division of General Internal Medicine.

## Footnotes

1 Question legend: 1. the report is that of a CXR, 2. the report definitely has no opacity suggesting pneumonia, 3. the report definitely has an opacity suggesting pneumonia, 4. the report is of a follow-up imaging report with minimal details in the text of the report, 5. the report shows the presence of an endotracheal tube.

